# Differences in the presentation and progression of Parkinson’s disease by sex

**DOI:** 10.1101/2020.04.08.20058370

**Authors:** Hirotaka Iwaki, Cornelis Blauwendraat, Hampton L. Leonard, Mary B. Makarious, Jonggeol J. Kim, Ganqiang Liu, Jodi Maple-Grødem, Jean-Christophe Corvol, Lasse Pihlstrøm, Marlies van Nimwegen, Luba Smolensky, Ninad Amondikar, Samantha J. Hutten, Mark Frasier, Khanh-Dung H. Nguyen, Jacqueline Rick, Shirley Eberly, Faraz Faghri, Peggy Auinger, Kirsten M. Scott, Ruwani Wijeyekoon, Vivianna M. Van Deerlin, Dena G. Hernandez, J. Raphael Gibbs, Aaron G. Day-Williams, Alexis Brice, Guido Alves, Alastair J. Noyce, Ole-Bjørn Tysnes, Jonathan R. Evans, David P. Breen, Karol Estrada, Claire E. Wegel, Fabrice Danjou, David K. Simon, Ole A. Andreassen, Bernard Ravina, Mathias Toft, Peter Heutink, Bastiaan R. Bloem, Daniel Weintraub, Roger A. Barker, Caroline H. Williams-Gray, Bart P. van de Warrenburg, Jacobus J. Van Hilten, Clemens R. Scherzer, Andrew B. Singleton, Mike A. Nalls

**Affiliations:** Laboratory of Neurogenetics, National Institute on Aging, National Institutes of Health, Bethesda, MD, USA; Data Tecnica International, Glen Echo, MD, USA; Advanced Center for Parkinson’s Disease Research, Brigham and Women’s Hospital and Harvard Medical School, Boston, MA, USA; Precision Neurology Program, Harvard Medical School, Brigham and Women’s Hospital, Boston, MA, USA; School of Medicine, Sun Yat-sen University, Guangzhou, Guangdong, China; The Norwegian Centre for Movement Disorders, Stavanger University Hospital, Stavanger, Norway; Department of Chemistry, Bioscience and Environmental Engineering, University in Stavanger, Stavanger, Norway; Assistance-Publique Hôpitaux de Paris, ICM, INSERM UMRS 1127, CNRS 7225, ICM, Department of Neurology and CIC Neurosciences, Pitié-Salpêtrière Hospital, Paris, France; Department of Neurology, Oslo University Hospital, Oslo, Norway; Department of Neurology, Donders Institute for Brain, Cognition, and Behaviour, Radboud University Medical Centre, Nijmegen, The Netherlands; The Michael J. Fox Foundation for Parkinson’s Research, New York, NY, USA; Translational Genome Sciences, Biogen, Cambridge, MA, USA; Department of Neurology University of Pennsylvania, Philadelphia, PA, USA; Department of Biostatistics and Computational Biology, University of Rochester, Rochester, NY, USA; Department of Neurology, Center for Health + Technology, University of Rochester, Rochester, NY, USA; Department of Clinical Neurosciences, University of Cambridge, John van Geest Centre for Brain Repair, Cambridge, UK; Department of Pathology and Laboratory Medicine, Center for Neurodegenerative Disease Research, Parelman School of Medicine at the University of Pennsylvania, Philadelphia, PA, USA; Flagship Labs 60 Inc, Cambridge, MA, USA; Statistical Genetics, Biogen, Cambridge, MA, USA; Institut du cerveau et de la moelle épinière ICM, Paris, France; Sorbonne Université SU, Paris, France; INSERM UMR1127, Paris, France; Department of Neurology, Stavanger University Hospital, Stavanger, Norway; Preventive Neurology Unit, Wolfson Institute of Preventive Medicine, Queen Mary University of London, London, UK; Department of Clinical and Movement Neurosciences, UCL Institute of Neurology, London, UK; Department of Neurology, Haukeland University Hospital, Bergen, Norway; Department of Clinical Medicine, University of Bergen, Bergen, Norway; Department of Neurology, Nottingham University NHS Trust, Nottingham, UK; Centre for Clinical Brain Sciences, University of Edinburgh, Edinburgh, Scotland; Anne Rowling Regenerative Neurology Clinic, University of Edinburgh, Edinburgh, Scotland; Usher Institute of Population Health Sciences and Informatics, University of Edinburgh, Edinburgh, Scotland; Department of Medical and Molecular Genetics, Indiana University, Indianapolis, IN, USA; Department of Neurology, Beth Israel Deaconess Medical Center, Boston, MA, USA; Harvard Medical School, Boston, MA, USA; NORMENT; Institute of Clinical Medicine, University of Oslo, Oslo, Norway, Norway; Division of Mental Health and Addiction, Oslo University Hospital, Oslo, Norway, Norway; Voyager Therapeutics, Cambridge, MA, USA; Department of Neurology, University of Rochester School of Medicine, Rochester, NY, USA; Institute of Clinical Medicine, University of Oslo, Oslo, Norway; German Center for Neurodegenerative Diseases-Tubingen, Tuebingen, Germany; HIH Tuebingen, Tubingen, Tuebingen, Germany; Department of Psychiatry, University of Pennsylvania School of Medicine, Philadelphia, PA, USA; Department of Veterans Affairs, Philadelphia, PA, USA; Department of Clinical Neurosciences and WT-MRC Cambridge Stem Cell Institute, University of Cambridge, Cambridge, UK; Department of Clinical Neurosciences, University of Cambridge, Cambridge, UK; Department of Neurology, Leiden University Medical Center, Leiden, The Netherlands

**Author notes:** **Corresponding author:** Mike A. Nalls Ph.D., Laboratory of Neurogenetics, National Institute on Aging, National Institutes of Health, 35 Convent Drive, Bethesda, MD 20892, USA.

## Abstract

**Objectives:** Identifying the contribution of biological sex to the heterogeneity in presentation and progression of Parkinson’s disease (PD).

**Background:** The different prevalence of Parkinson’s disease (PD) in men and women suggests that sex-associated mechanisms influence disease mechanisms. Investigating the contribution of sex to disease heterogeneity may uncover disease processes, and lead to new therapeutic targets. Also, understanding these differences in phenotypes will result in better patient management and the planning of more efficient clinical trials.

**Methods:** We tested 40 clinical phenotypes using longitudinal clinic-based patient cohorts consisting of 5,946 patients with a median follow-up of 3.1 years. For continuous outcomes, we used linear regressions at baseline to test the sex-associated differences in presentation, and linear mixed-effects models to test the sex-associated differences in progression. For binomial outcomes, we used logistic regression models at baseline and Cox models for survival analyses. We adjusted for age, disease duration and dopaminergic medication usage. In the secondary analyses, data from 28,809 PD patients and 10,556 non-PD participants from Fox Insight, an online-only self-assessment cohort for PD research, were analyzed to check whether the sex-associated differences observed in the primary analyses were consistent in the cohort and whether the differences were unique to PD or not.

**Results:** Female PD patients had a higher risk for developing dyskinesia early during the follow-up period; with a slower progression in their difficulties of activities of daily living as measured by the Unified Parkinson’s Disease Rating Scale Part II (classic/MDS-revised version); and a lower risk of developing cognitive impairment than male patients. The findings in the longitudinal clinic-based cohorts were mostly consistent with the results in the online-only cohort.

**Conclusions:** This large-scale analysis observed the sex contribution to the heterogeneity of Parkinson’s disease. The results highlight the necessity of future research of the underlying mechanism and importance of personalized clinical management.

## Introduction

The prevalence of Parkinson’s disease (PD) is 1.5 −2.0 times higher in men than in women. This discrepancy suggests that there be sex-associated factors that modify the disease process. Identifying the interplay between sex and the disease has the potential to develop disease-modifying therapy as well as to inform patient management and the planning of more efficient clinical trials. Researchers have investigated the sex-associated differences in phenotypes among patients with PD.^1^ Male PD patients were reported to have more akinesia/rigid features,^2^ cognitive impairment,^3,4^ and rapid eye movement sleep behavioral disorder.^5,6^ On the other hand, depression,^7–10^ and dyskinesia ^7,11–13^ were documented to occur more frequently in female patients. However, these studies were generally small in sample size and predominantly done in a cross-sectional setting.

In this work, we analyzed longitudinal data from 12 PD cohorts representing 5,946 participants with a median of 3.7 years of follow-up. There were two objectives: (1) to identify the baseline differences between men and women in disease presentation and (2) to identify the influence of sex on longitudinal symptom trajectory. Further, we analyzed the Fox Insight dataset, an online-only Parkinson’s disease research cohort, to assess whether the observations in the longitudinal datasets were consistent in an independent dataset. Moreover, analyzing the data from both PD participants and non-PD participants of the Fox Insight dataset provided the opportunity to evaluate if there were any differences in the prevalence of self-reported outcomes between participants with and without PD. This analysis further illustrated that some of the differences may be inherited from general differences between males and females, while others are disease-specific.

## Methods

### Participants

#### 12 longitudinal cohorts

We analyzed data from 12 longitudinal PD cohorts from North America, Europe, and Australia in this study (Table 1). Among them, the following 4 studies enrolled people with early phase PD not being treated at study enrollment (de novo cohorts): Parkinson’s Progression Markers Initiative (PPMI), Parkinson Research Examination of CEP□1347 Trial study with its subsequent prospective study (PreCEPT/PostCEPT), the Norwegian ParkWest study (PARKWEST), and Deprenyl and Tocopherol Antioxidative Therapy of Parkinsonism (DATATOP). Other cohorts included Parkinsonism Incidence and Cognitive and Non□motor heterogeneity In Cambridgeshire (PICNICS), National Institutes of Health Exploratory Trials in Parkinson’s Disease Large Simple Study 1 (NET_PD_LS1), Drug Interaction With Genes in Parkinson’s Disease (DIGPD), Parkinson’s Disease Biomarker Program (PDBP), Harvard Biomarkers Study (HBS), ParkFit Study (PARKFIT), Profiling Parkinson’s Disease Study (PROPARK), and Udall Centers program (UDALL_PENN). Participants’ information was obtained under appropriate written consent and with local institutional and ethical approval. The summary of the designs and inclusion/exclusion criteria of these cohorts are documented in Supplemental Materials.

**Table 1.** Baseline characteristics of study cohorts.

| Cohort | N | f-u, year | Female, % | Age, year old | European % | Duration, y | LD,<br>% | DA,<br>% |
| --- | --- | --- | --- | --- | --- | --- | --- | --- |
| PPMI | 408 | 7.0 | 34.6 | 61.67 (9.78) | 95.1 | 0.55 | - | - |
| PreCEPT_PostCEPT | 390 | 6.9 | 33.8 | 60.21 (9.53) | 97.7 | 0.81 | - | - |
| PARKWEST | 181 | 5.0 | 37.8 | 67.94 (9.18) | 100 | 0.18 | - | - |
| DATATOP | 796 | 0.8 | 33.7 | 61.08 (9.51) | 97.7 | 1.14 | - | - |
| PICNICS | 122 | 2.1 | 35.2 | 67.88 (9.07) | 98.4 | 4.17 | 29.5 | 19.7 |
| NET_PD_LS1 | 1705 | 3.0 | 35.7 | 61.76 (9.60) | 92.7 | 1.55 | 56.7 | 61.4 |
| DIGPD | 350 | 2.0 | 39.4 | 61.83 (10.05) | 85.8 | 2.51 | 64.3 | 72 |
| PDBP | 486 | 2.8 | 39.7 | 64.97 (8.94) | 93.0 | 5.27 | 80 | 52.8 |
| HBS | 482 | 1.8 | 35.3 | 66.08 (9.57) | 96.3 | 4.17 | 72.4 | 40.5 |
| PROPARK | 327 | 4.7 | 33.9 | 59.54 (10.74) | NA | 6.57 | 66.1 | 73.1 |
| PARKFIT | 466 | 2.0 | 33.3 | 65.35 (7.47) | NA | 5.10 | NA | NA |
| UDALL_PENN | 233 | 4.1 | 30.9 | 70.41 (7.55) | 94.4 | 6.01 | 86.3 | 49.4 |
| Fox Insight (nonPD) | 7588 | - | 78.8 | 62.73 (7.51) | 95.8 | - | - | - |
| Fox Insight (PD) | 17719 | - | 45 | 66.39 (7.14) | 96.4 | 4.56 | 78.8 | 31.9 |

f-u, median follow-up period; European, European descent; Duration, mean disease duration; LD, levedopa use; DA, dopamine agonist use. Age, mean (standard deviation).
DATATOP, Deprenyl and Tocopherol Antioxidative Therapy of Parkinsonism; DIGPD, Drug Interaction with Genes in Parkinson's Disease; HBS, Harvard Biomarkers Study; NET-PD\_LS1, NIH Exploratory Trials in Parkinson's Disease Large Simple Study 1; PARKFIT, ParkFit study; PARKWEST, The Norwegian ParkWest study; PDBP, Parkinson's Disease Biomarker Program; PICNICS, Parkinsonism Incidence and Cognitive and Non-motor heterogeneity In Cambridgeshire; PPMI, Parkinson's progression markers initiative; PreCEPT\_PostCEPT, Parkinson Research Examination of CEP-1347 Trial and PostCEPT; PROPARK, Profiling Parkinson's disease study; and UDALL\_PENN, Morris K. Udall Centers for Parkinson's Research.

#### Fox Insight

To evaluate the consistency of results from the longitudinal dataset, we explored the independent dataset, Fox Insight. Fox insight is an online-only Parkinson’s disease research cohort.^14^ Details of the study are available online (https://foxinsight.michaeljfox.org/). Individuals aged 18 or older with or without PD were enrolled through in-person referral or online advertisements. The participants provided online informed consent, and self-reported demographic, characteristics, symptoms, medical history, and PD medication were collected. This is a longitudinal study, but we analyzed the data cross-sectionally for the current work because follow-up periods were relatively short (e.g. the median follow-up period was 0.4 years for Non Motor Symptoms Questionnaire). In the analysis step, we adjusted for age and disease duration and to limit the impact of the extreme data points, we included participants from the middle 80% of the age distribution and disease duration distribution (only for PD participants). This means that we excluded the participants aged less than the lower 10th percentile (<46.8 years old) or more than the 90th percentile (>77.4 years old), and PD patients with a disease duration less than 1 year (10th percentile) and more than 13.5 years (90th percentile).

### Measurements

#### Clinical Data Harmonization of 12 cohorts

Twenty three measurements, 11 binomial and 9 continuous measurements, were analyzed as outcome measures. Binomial outcomes included constipation, mild cognitive impairment, depression, daytime sleepiness, hyposmia, insomnia, wearing off, dyskinesias, RBD, restless legs syndrome, and modified Schwab and England Activities of Daily Living Scale scores of 70 or less (SEADL70). Some binomial outcomes had study-specific outcomes and these criteria are summarized in Supplemental Materials. For continuous outcomes, we collected Hoehn and Yahr (HY) stage scale, total and subLscores of the Unified Parkinson’s Disease Rating Scale (UPDRS) or the Movement Disorder Society–revised version (MDS□UPDRS), Mini□Mental State Examination, Montreal Cognitive Assessment (MoCA), and modified Schwab and England Activities of Daily Living Scale (SEADL). UPDRS scores were normalized to the z values (UPDRS*_scaled).

#### Fox Insight

The February 2020 data cut was downloaded from https://foxden.michaeljfox.org.^15^ The demographic and disease status were obtained from enrollment and registration questionnaires. For clinical outcomes of interest, we obtained the responses from the following questionnaires: Geriatric Depression Scale (GDS) for depression (score 6 or more),^16^ Non-Motor Symptoms Questionnaire (NMS-QUEST) for constipation, depressed mood (Mood depressed) and proxy for lack of the sense of smell/taste,^17^ MDS-UPDRS Part II questionnaire, REM Sleep Behaviour Disorder Single-Question Screen,^18^ 15-item Penn Parkinson’s Disease Daily Activities Questionnaire (PDAQ-15) for cognition-related instrumental functional abilities,^19^ and Understanding the impact of Off and On in Parkinson’s patients Questionnaire for dyskinesia and wearing off.

### Statistical analysis

Linear and logistic models were used to analyze the baseline difference in the presentation of PD between male and female patients per cohort. For binomial outcomes, 25 or more outcomes should be observed in the cohort to be analyzed. Covariates were the linear and square terms of age and disease duration to adjust for the linear and non-linear effects. In addition, we adjusted for levodopa and dopamine agonist usage. To test the differences in the rates of progression in continuous outcomes, we used linear mixed-effects models with the same covariates as the baseline models and with the random effects on individual intercept and slope (change per year). We evaluated sex-associated differences in the rate of change by testing the interaction between sex and years in study. The survival analyses were conducted among those who did not have an outcome at baseline. Cox regression models were used adjusting for the same covariates as those in the baseline models. The outcomes with less than 20 events over the follow-up period were not analyzed. The R model statements for these analyses were summarized in Supplemental Materials.

Then, we combined the cohort-level results with an inverse variance weighted random effect model. We focused on the robust associations throughout the cohorts so the meta-analyses with p-values less than 0.05 for a test of homogeneity were excluded from further evaluations. The association with the two-sided p-value of 0.05 after Bonferroni-correction for the number of total analyses were considered significant.

For the analysis of the Fox Insight dataset, we tested two terms: the mean difference between males and females (main term) and the interaction between sex and disease duration (interaction term). The adjusted covariates were linear and square age, linear and square disease duration, and indicators for levodopa and dopamine agonist usage. We further analyzed the association with sex and outcomes among non-PD participants, adjusted for linear and square age. Then we conducted the test of homogeneity between these sex-associated differences among PD cases and non-PD participants to evaluate whether the sex differences were PD-specific or reflected differences seen in the non-PD population. In the analyses for this dataset, we used a significance level of 0.05 for the raw p-value because the purpose of the analyses was to evaluate consistency with the longitudinal analyses.

All the statistical analyses and drawings were conducted using R version 3.6 and python version 3.7. The analysis scripts are available at https://github.com/neurogenetics/PDpheno_by_sex.

## Results

The cohort participants are summarized in Table 1. Participants in these cohorts were of different ages and in different stages of Parkinson’s disease, but overall they were in relatively early phases of their disease. The majority of the participants were of European descent. Interestingly, Fox Insight had more female participants than other cohorts and the ratio of females to males was especially high among non-PD participants in Fox Insight as previously described.^20^

In total, we conducted 40 meta-analyses using the clinic-based longitudinal data, of which 3 were rejected by a test of heterogeneity with a significance level of 0.05. With the Bonferroni correction of multiple comparisons, we set our p-value (P) threshold as 0.05/37 =0.00135. Among these associations, 9 were significant and the direction and magnitude of associations of being female against being male are shown in Table 2 and Figure 1. (All meta-analysis results are in Supplemental Materials).

**Table 2.**
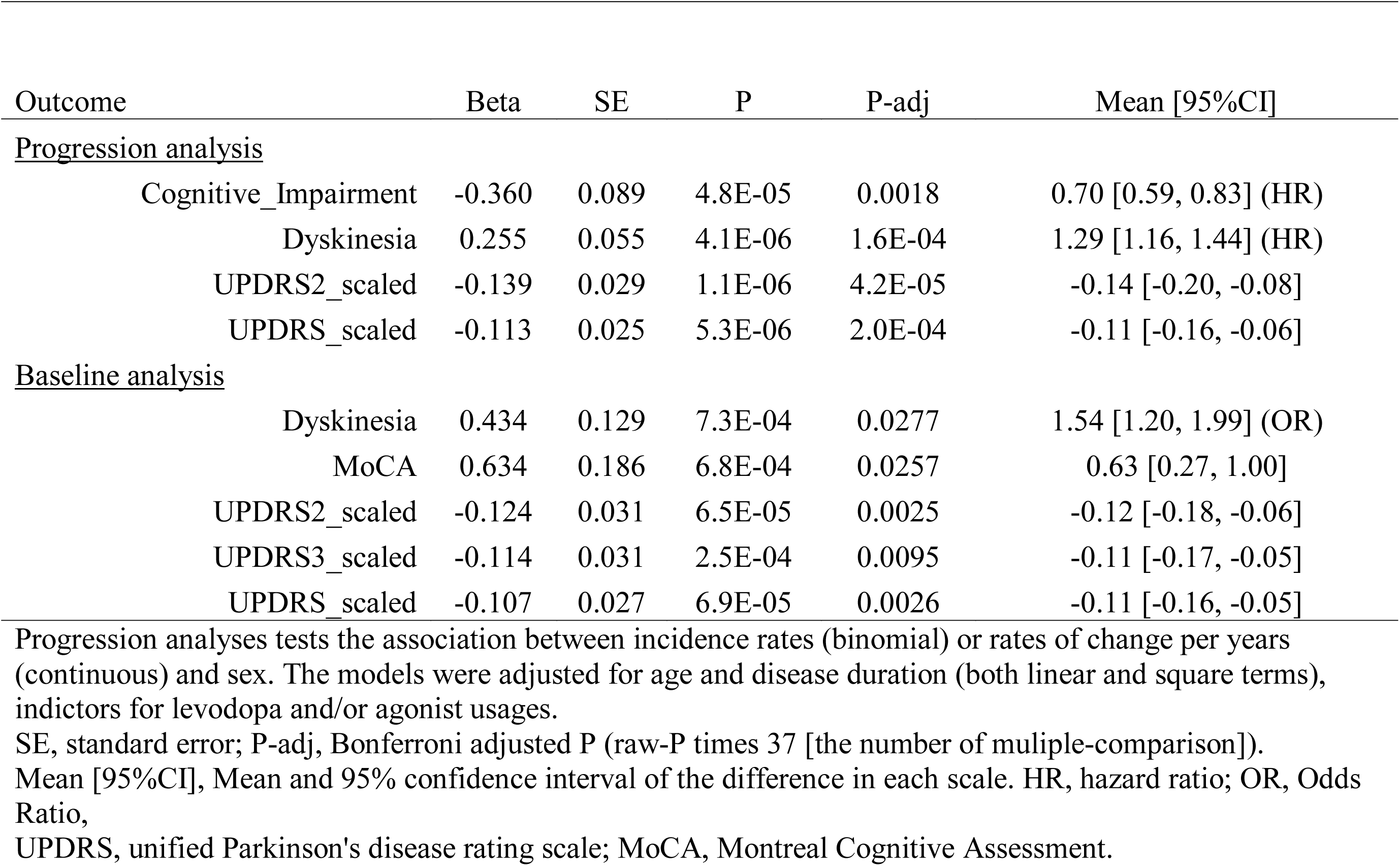
Meta-analysis results for significant associations with sex and phenotypes.

| Outcome | Beta | SE | P | P-adj | Mean [95%CI] |
| --- | --- | --- | --- | --- | --- |
| <u>Progression analysis</u> |  |  |  |  |  |
| Cognitive_Impairment | -0.360 | 0.089 | 4.8E-05 | 0.0018 | 0.70 [0.59, 0.83] (HR) |
| Dyskinesia | 0.255 | 0.055 | 4.1E-06 | 1.6E-04 | 1.29 [1.16, 1.44] (HR) |
| UPDRS2_scaled | -0.139 | 0.029 | 1.1E-06 | 4.2E-05 | -0.14 [-0.20, -0.08] |
| UPDRS_scaled | -0.113 | 0.025 | 5.3E-06 | 2.0E-04 | -0.11 [-0.16, -0.06] |
| <u>Baseline analysis</u> |  |  |  |  |  |
| Dyskinesia | 0.434 | 0.129 | 7.3E-04 | 0.0277 | 1.54 [1.20, 1.99] (OR) |
| MoCA | 0.634 | 0.186 | 6.8E-04 | 0.0257 | 0.63 [0.27, 1.00] |
| UPDRS2_scaled | -0.124 | 0.031 | 6.5E-05 | 0.0025 | -0.12 [-0.18, -0.06] |
| UPDRS3_scaled | -0.114 | 0.031 | 2.5E-04 | 0.0095 | -0.11 [-0.17, -0.05] |
| UPDRS_scaled | -0.107 | 0.027 | 6.9E-05 | 0.0026 | -0.11 [-0.16, -0.05] |

Progression analyses tests the association between incidence rates (binomial) or rates of change per years (continuous) and sex. The models were adjusted for age and disease duration (both linear and square terms), indicators for levodopa and/or agonist usages.
SE, standard error; P-adj, Bonferroni adjusted P (raw-P times 37 [the number of multiple-comparison]).
Mean [95%CI], Mean and 95% confidence interval of the difference in each scale. HR, hazard ratio; OR, Odds Ratio,
UPDRS, unified Parkinson's disease rating scale; MoCA, Montreal Cognitive Assessment.

**Figure 1:**
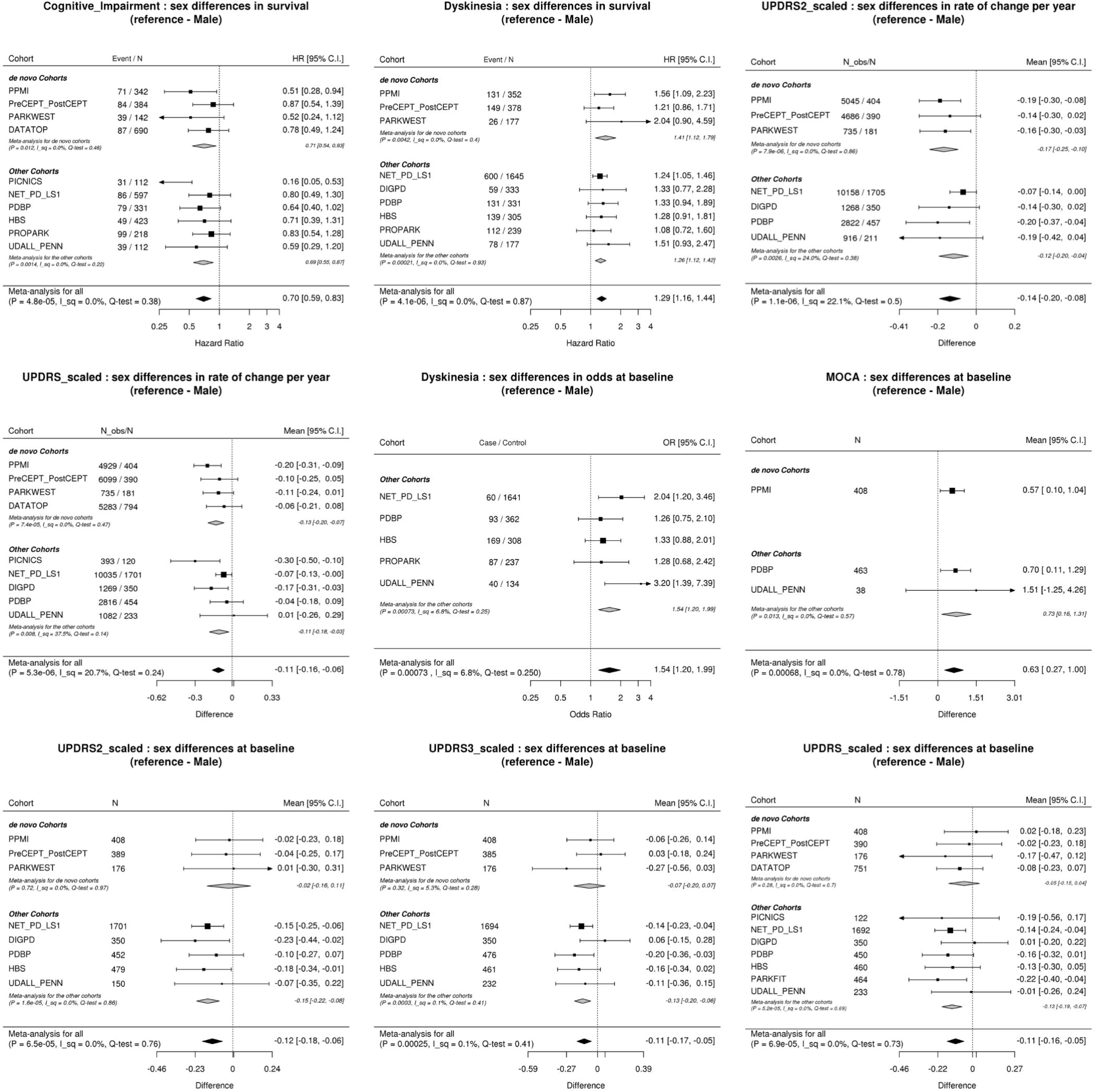
The forest plots for the sex differences in outcomes. DATATOP, Deprenyl and Tocopherol Antioxidative Therapy of Parkinsonism; DIGPD, Drug Interaction with Genes in Parkinson’s Disease; HBS, Harvard Biomarkers Study; NET-PD_LS1, NIH Exploratory Trials in Parkinson’s Disease Large Simple Study 1; PARKFIT, ParkFit study; PARKWEST, The Norwegian ParkWest study; PDBP, Parkinson’s Disease Biomarker Program; PICNICS, Parkinsonism Incidence and Cognitive and Non-motor heterogeneity In Cambridgeshire; PPMI, Parkinson’s progression markers initiative; PreCEPT_PostCEPT, Parkinson Research Examination of CEP-1347 Trial and PostCEPT; PROPARK, Profiling Parkinson’s disease study; and UDALL_PENN, Morris K. Udall Centers for Parkinson’s Research. P, non-adjusted p-values; I_sq, I^2^ statistic; QEp, test of heterogeneity.

Female PD patients were less likely to develop cognitive impairment over time (HR 0.70 [0.59, 0.83], P = 4.8E-5). The stronger association was observed when we adjusted for years of education. (HR 0.63 [0.53, 0.76], P = 4.3E-7, Supplemental Material). Also, the baseline MoCA scores were higher in female patients (0.63 [0.27, 1.00]) while baseline MMSE score was not significantly different between sexes (P = 0.97, Supplemental Materials).

Female patients had a higher rate of developing dyskinesia (HR 1.29 [1.16, 1.44]). To assess the impact of weight, BMI and medication on the association, we conducted ad-hoc analyses on a subset of data (PDBP, PPMI and NET_PD_LS1. 2281 participants) where height at baseline, weight at baseline, and medication at visits were recorded. We adjusted analyses for each of these factors. With weight adjustment the association was no longer significant (P = 0.059), while the magnitude of the association became larger when adjusted for levodopa dosages or levodopa equivalent dosages.

Adjusting for BMI did not substantially change the magnitude of the association (Beta: from 0.285 to 0.250) and the sex difference was still significant. (Supplemental Materials). Consistent with the higher incidence rate of dyskinesia in female patients, prevalent female PD patients in non de novo cohorts also had more dyskinesia at baseline.

Activities of daily living, captured in the UPDRS Part II score, was better in female PD patients in the baseline analysis (--0.12 [−0.18, −0.06] in z score), and the progression rate was less in female patients (−0.14 [−0.20, −0.08] in z score per year). A more detailed analysis of the forest plots of the UPDRS Part II scores at baseline showed that the associations between sex and UPDRS Part II were not apparent among the de novo cohorts but rather driven by the differences observed in the non-de novo cohorts(Figure 1). While we did not find significant sex-associated differences in progression rate in UPDRS Part I/III/IV, the rate of change in the total scores of UPDRS were significantly milder in female patients (−0.11 [0.16, −0.06] per year in z score). When only considering the de novo cohorts, they reported similar results-progression rate was slower in female patients (−0.14 [−0.21, −0.07] in z score per year, P = 2.6E-5, Supplemental Materials).

Finally, female patients also had lower scores on the UPDRS Part III and the UPDRS total score compared to males in the baseline analyses.

In the analyses of similar phenotypes within the Fox Insight dataset (Table 3), we generally confirmed the results of the longitudinal dataset analyses. In the Fox Insight dataset analysis, the interaction terms between sex and disease duration indicated the average sex-associated differences in the longitudinal trajectories for the outcomes. For example, the positive association with the interaction and PDAQ-15 meant that, in the Fox Insight dataset, the scores of PDAQ-15 in female patients when compared with that in males were higher (i.e., better cognition-related instrumental functional abilities) among patients with longer disease durations. To illustrate this, we visualized the sex differences stratified by disease duration (supplemental materials). The result is consistent with that of the longitudinal dataset analysis that female patients had a lower risk of developing cognitive impairment during the course. Similarly, the results from the Fox Insight dataset were consistent with a higher rate of developing dyskinesia; and the lower scores and a slower deterioration rate in UPDRS Part II in female patients, as observed in the longitudinal analyses.

**Table 3.**
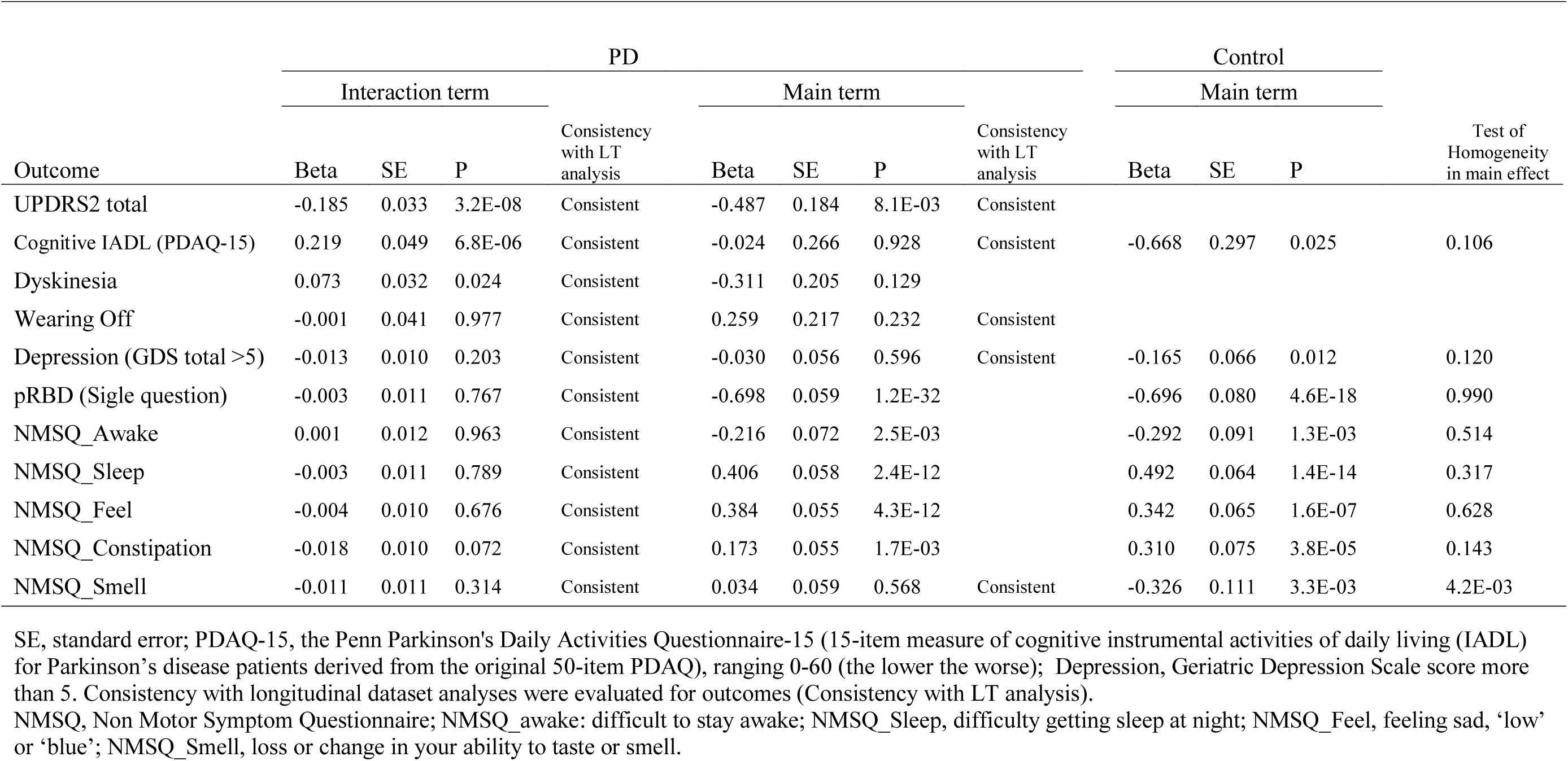
Analysis results for sex difference in main term and interaction term with sex and disease duration in replication cohort.

In addition, null differences between male and female patients in presentation and progression of wearing off, depression, and hyposmia were also supported in the Fox Insight dataset. In contrast, loss of the sense of smell/taste was significantly more frequently reported in males in the controls. Having PD might diminish the general sex difference in this phenotype. Single question answers for RBD and some NMSQuest questionnaire regarding “difficult to stay awake” (NMSQ_awake), “difficulty in getting to sleep” (NMSQ_Sleep), “feeling sad, low or blue” (NMSQ_feel), and NMSQ_Constipation were significantly different by sex in the Fox Insight dataset. The prevalence of similar outcomes such as pRBD, daytime sleepiness, insomnia, depression and constipation were not significantly associated with sex in the meta-analyses of 12 longitudinal cohorts. However, the test for these associations give raw p-values less than 0.05 with the same directions as the Fox Insight results. So it may be that there are sex-phenotype associations but the primary analyses didn’t have a large enough sample size to detect these. Of note, all of these sex-phenotype associations among PD participants in Fox Insight were also significant among non-PD participants. In addition, based on the test of homogeneity between the results from PD and non-PD participants, there was no evidence that the magnitude of these associations were different between PD and non-PD participants.

## Discussion

We analyzed clinic-based longitudinal data from 5,946 participants and meta-analyzed the difference in presentation and progression of phenotypes between men and women with PD. We also used web-based online cohorts and analyzed data from 28,809 PD patients and 10,556 non-PD to confirm our results. The current results suggest that female PD patients develop dyskinesia early, progress more slowly with respect to activities of daily living (ADL) restrictions and are less likely to develop cognitive impairment. For some non-motor symptoms asked in online questionnaires (such as pRBD, daytime sleepiness, insomnia, depressive mood and constipation), we found significant sex-associated differences among PD participants only in the Fox Insight dataset, but we could not identify clear PD-specific differences when comparing PD and non-PD participants for these associations.

Some studies have previously reported that female patients had more risk of developing earlier and more severe dyskinesia,^7,11^ and longer duration of dyskinesia.^12^. This is consistent with the faster development of dyskinesia among female patients and a greater rate of increase in UPDRS Part IV scores in our study. The reason for this phenomenon is not fully understood, but relatively higher levodopa dosages according to body weight in females may be partially responsible.^13^ Our ad-hoc analyses also suggested the importance of body weight on the association between sex and early development of dyskinesia.

Regarding the impact on activities of daily living, contradictory results have been reported previously. Two studies evaluated patients who undertook the surgical treatment: one study observed no difference in UPDRS Part II between males and females, while the other reported females had worse scores. In their studies, females had a longer duration of the disease, which may have affected the results. Another cross-sectional study also reported worse UPDRS Part II in female patients.^7^ They reported that among 5 categories of overall ADL capacity, having the most severe 2 categories was more frequent in females using a chi-square test while our analyses used UPDRS Part II and multiple regression models. These different outcome measurements and statistical approaches may account for these different results.

The slower development of cognitive decline in female patients was reported from some longitudinal studies.^3,4,21^ The executive and attention features were primarily affected in PD. We observed the discrepancy between MoCA and MMSE in the baseline analyses and this discrepancy may be partly because MoCA is more sensitive in detecting executive dysfunction than MMSE.^22^. In contrast, the longitudinal differences in the rates of decline in MoCA or MMSE were not significantly different by sex in our data. Interestingly, MoCA scores were sometimes reported to be higher in healthy aging women than men.^23–25^ The slower development of cognitive impairment in females may in part reflect their relative high baseline ability in the areas susceptible for Parkinson’s disease.

Several associations reported previously were not observed in the current data. pRBD was reported to be more prevalent in males with PD,^5,6^ while some disagree.^26,27^ We, too, were unable to confirm the association in the current longitudinal dataset. Although the prevalence of pRBD detected by single question screening was higher in male patients in the Fox Insight dataset, the similarly higher prevalence of pRBD in non-PD male participants raises questions as to the PD-specific nature of this association. Female PD patients were more depressed according to previous reports.^7–10^ We did not confirm this sex-associated difference in presentation and progression in our longitudinal data or the Fox Insight dataset. However, female PD patients expressed a depressive mood more frequently than male patients in response to the related NMSQuest question (‘feeling sad, ‘low’ or ‘blue’) in the Fox Insight dataset. But again, the magnitude of the associations was not different between PD and non-PD participants, indicating that the sex difference in this outcome may be not PD-specific.

There are some limitations to the current work. Fox Insight is an online-only cohort and inherently different from a clinic-based cohort; however, our analyses were mostly consistent across these two different settings. Also, because the study participants were almost all of European descent, it will be important to test the generalizability of these observations across different ancestrally distinct groups. In this study, we focused on the overall associations between sex and phenotypes and did not separate the biological mechanisms from environmental mechanisms. We believe that it will be important to examine the potential effects of environmental factors such as estrogen usage, history of pregnancy, tobacco use, and pesticide exposure, that may contribute to the differences between male patients and female patients.

Despite some limitations, the current work has some important meanings. First, the total number of participants in our longitudinal analysis was one of the largest studied. Second, although each study had different cohort characteristics, we controlled for heterogeneity and multiple comparisons to detect robust signals. Indeed, most of these associations between sex and disease presentation/progression were consistent with analyses in the independent Fox Insight dataset. Third, our results could be generalized to PD patients across various disease stages in different contexts given the range of studies incorporated. Fourth, by comparing PD with non-PD individuals, we obtained an insight into whether sex-associated phenotypes in PD were disease-specific or rather reflecting general differences by sex. Finally, female PD patients have been an underrepresented population in clinical trials.^2^ The current work emphasizes the importance of recognizing gender bias in developing a treatment for Parkinson’s disease in the real world.

In conclusion, we observed that female PD patients were earlier in developing dyskinesia in their disease course and progressed more slowly with respect to cognitive deficits and problems with ADL compared to male PD patients. The associations were mostly consistent across the different longitudinal cohorts and the online survey.

## Data Availability

Qualified investigators can request raw data through the organizations' homepages (PDBP: https://pdbp.ninds.nih.gov/, PPMI: https://www.ppmi-info.org/, Fox Insight:https://foxinsight.michaeljfox.org/) or collaboration.

## Acknowledgement

We thank all study participants and their family, investigators and members of the following Studies: Parkinson Study Group: Deprenyl and Tocopherol Antioxidative Therapy of Parkinsonism (DATATOP); Drug Interaction with Genes in Parkinson’s Disease (DIGPD); Harvard Biomarkers Study (HBS); NET-PD_LS1, NIH Exploratory Trials in Parkinson’s Disease Large Simple Study 1; The Norwegian ParkWest study (ParkWest); Parkinson’s Disease Biomarker Program (PDBP);Parkinsonism Incidence and Cognitive and Non-motor heterogeneity In Cambridgeshire (PICNICS); Parkinson’s progression markers initiative (PPMI); Parkinson Study Group: Parkinson Research Examination of CEP-1347 Trial (PreCEPT) and its following study (PostCEPT); Profiling Parkinson’s disease study (ProPark); Morris K. Udall Centers for Parkinson’s Research (Udall); and Fox Insight study.

PPMI – a public-private partnership – is funded by The Michael J. Fox Foundation for Parkinson’s Research and funding partners, including AbbVie, Allergan, Avid Radiopharmaceuticals, Biogen, BioLegend, Bristol-Myers Squibb, Celgene, Denali Incorporated, GE Healthcare, Genentech, GlaxoSmithKline, Eli Lilly and Company, Lundbeck, Merck & Co., Meso Scale Discovery, Pfizer, Piramal, Prevail Therapeutics, Roche, Sanofi Genzyme, Servier Laboratories, Takeda, Teva, UCB, Verily, Voyager Therapeutics, and Golub Capital (www.ppmi-info.org/fundingpartners). The Fox Insight Study (FI) is funded by The Michael J. Fox Foundation for Parkinson’s Research. We would like to thank the Parkinson’s community for participating in this study to make this research possible.

We also thank the following grants and financial supporters of above studies; DATATOP was supported by a Public Health Service grant (NS24778) from the National Institute of Neurological Disorders and Stroke (NINDS); by grants from the General Clinical Research Centers Program of the National Institutes of Health at Columbia University (RR00645), the University of Virginia (RR00847), the University of Pennsylvania (RR00040), the University of Iowa (RR00059), Ohio State University (RR00034), Massachusetts General Hospital (RR01066), the University of Rochester (RR00044), Brown University (RR02038), Oregon Health Sciences University (RR00334), Baylor College of Medicine (RR00350), the University of California (RR00827), Johns Hopkins University (RR00035), the University of Michigan (RR00042), and Washington University (RR00036), the Parkinson’s Disease Foundation at Columbia-Presbyterian Medical Center, the National Parkinson Foundation, the Parkinson Foundation of Canada, the United Parkinson Foundation, Chicago, the American Parkinson’s Disease Association, New York, and the University of Rochester; DIGPD is supported by Assistance Publique Hôpitaux de Paris, funded by a grant from the French Ministry of Health (PHRC 2008, AOM08010) and a grant from the Agence Nationale pour la Sécurité des Médicaments (ANSM 2013); HBS is supported by the Harvard NeuroDiscovery Center, Michael J Fox Foundation, NINDS U01NS082157, U01NS100603, and the Massachusetts Alzheimer’s Disease Research Center NIA P50AG005134; NET-PD_LS1 was supported by NINDS grants U01NS043128; ParkFit is supported by ZonMw (the Netherlands Organization for Health Research and Development (75020012)) and the Michael J Fox Foundation for Parkinson’s research, VGZ (health insurance company), GlaxoSmithKline, and the National Parkinson Foundation; ParkWest is supported by the Research Council of Norway, the Western Norway Regional Health Authority, Stavanger University Hospital Research Funds, and the Norwegian Parkinson’s Disease Association; PDBP is a consortium with NINDS initiative; PICNICS has received funding from the Cure Parkinson’s Trust, the Van Geest Foundation and is supported by the National Institute of Health Research Cambridge Biomedical Research Centre; PPMI is supported by the Michael J Fox Foundation for Parkinson’s research; PreCEPT and PostCEPT were funded by NINDS 5U01NS050095□05, Department of Defense Neurotoxin Exposure Treatment Parkinson’s Research Program. Grant Number: W23RRYX7022N606, the Michael J Fox Foundation for Parkinson’s research, Parkinson’s Disease Foundation, Lundbeck Pharmaceuticals. Cephalon Inc, Lundbeck Inc, John Blume Foundation, Smart Family Foundation, RJG Foundation, Kinetics Foundation, National Parkinson Foundation, Amarin Neuroscience LTD, CHDI Foundation Inc, National Institutes of Health (NHGRI, NINDS), Columbia Parkinson’s Disease Research Center; ProPARK is funded by the Alkemade-Keuls Foundation, Stichting Parkinson Fonds, Parkinson Vereniging, The Netherlands Organisation for Health Research and Development; Udall is supported by NINDS. DPB is funded by a Wellcome Clinical Research Career Development Fellowship.

## References

1. Miller, I. N. & Cronin-Golomb, A. Gender differences in Parkinson’s disease: clinical characteristics and cognition. Mov. Disord. 25, 2695–2703 (2010).

2. Haaxma, C. A. et al.. Gender differences in Parkinson’s disease. J. Neurol. Neurosurg. Psychiatry 78, 819–824 (2007).

3. Locascio, J. J., Corkin, S. & Growdon, J. H. Relation between clinical characteristics of Parkinson’s disease and cognitive decline. J. Clin. Exp. Neuropsychol. 25, 94–109 (2003).

4. Cholerton, B. et al.. Sex differences in progression to mild cognitive impairment and dementia in Parkinson’s disease. Parkinsonism Relat. Disord. 50, 29–36 (2018).

5. Ozekmekçi, S., Apaydin, H. & Kiliç, E. Clinical features of 35 patients with Parkinson’s disease displaying REM behavior disorder. Clin. Neurol. Neurosurg. 107, 306–309 (2005).

6. Yoritaka, A., Ohizumi, H., Tanaka, S. & Hattori, N. Parkinson’s disease with and without REM sleep behaviour disorder: are there any clinical differences? Eur. Neurol. 61, 164–170 (2009).

7. Baba, Y., Putzke, J. D., Whaley, N. R., Wszolek, Z. K. & Uitti, R. J. Gender and the Parkinson’s disease phenotype. J. Neurol. 252, 1201–1205 (2005).

8. Fernandez, H. H., Lapane, K. L., Ott, B. R. & Friedman, J. H. Gender differences in the frequency and treatment of behavior problems in Parkinson’s disease. SAGE Study Group. Systematic Assessment and Geriatric drug use via Epidemiology. Mov. Disord. 15, 490–496 (2000).

9. Riedel, O. et al.. Cognitive impairment in 873 patients with idiopathic Parkinson’s disease. Results from the German Study on Epidemiology of Parkinson’s Disease with Dementia (GEPAD). J. Neurol. 255, 255–264 (2008).

10. Scott, B., Borgman, A., Engler, H., Johnels, B. & Aquilonius, S. M. Gender differences in Parkinson’s disease symptom profile. Acta Neurol. Scand. 102, 37–43 (2000).

11. Accolla, E. et al.. Gender differences in patients with Parkinson’s disease treated with subthalamic deep brain stimulation. Mov. Disord. 22, 1150–1156 (2007).

12. Hariz, G.-M., Lindberg, M., Hariz, M. I. & Bergenheim, A. T. Gender differences in disability and health-related quality of life in patients with Parkinson’s disease treated with stereotactic surgery. Acta Neurol. Scand. 108, 28–37 (2003).

13. Zappia, M. et al.. Body weight influences pharmacokinetics of levodopa in Parkinson’s

14. Smolensky, L. et al.. Fox Insight collects online, longitudinal patient-reported outcomes and genetic data on Parkinson’s disease. Scientific Data 7, 1–9 (2020).

15. Fox Login Page. https://doi.org/10.25549/bxya-6133.

16. Yesavage, J. A. et al.. Development and validation of a geriatric depression screening scale: A preliminary report. J. Psychiatr. Res. 17, 37–49 (1982).

17. Chaudhuri, K. R. et al.. International multicenter pilot study of the first comprehensive self-completed nonmotor symptoms questionnaire for Parkinson’s disease: the NMSQuest study. Mov. Disord. 21, 916–923 (2006).

18. Postuma, R. B. et al.. A single-question screen for rapid eye movement sleep behavior disorder: a multicenter validation study. Mov. Disord. 27, 913–916 (2012).

19. Brennan, L. et al.. The Penn Parkinson’s Daily Activities Questionnaire-15: Psychometric properties of a brief assessment of cognitive instrumental activities of daily living in Parkinson’s disease. Parkinsonism Relat. Disord. 25, 21–26 (2016).

20. Chahine, L. M. et al.. Comparison of an Online-Only Parkinson’s Disease Research Cohort to Cohorts Assessed In Person. J. Parkinsons. Dis. (2020) doi:10.3233/JPD-191808.

21. Pigott, K. et al.. Longitudinal study of normal cognition in Parkinson disease. Neurology 85, 1276–1282 (2015).

22. Pendlebury, S. T. et al.. Differences in cognitive profile between TIA, stroke and elderly memory research subjects: a comparison of the MMSE and MoCA. Cerebrovasc. Dis. 34, 48–54 (2012).

23. Thomann, A. E. et al.. The Montreal Cognitive Assessment: Normative Data from a German-Speaking Cohort and Comparison with International Normative Samples. J. Alzheimers. Dis. 64, 643–655 (2018).

24. Konstantopoulos, K., Vogazianos, P. & Doskas, T. Normative Data of the Montreal Cognitive Assessment in the Greek Population and Parkinsonian Dementia. Arch. Clin. Neuropsychol. 31, 246–253 (2016).

25. Borland, E. et al.. The Montreal Cognitive Assessment: Normative Data from a Large Swedish Population-Based Cohort. J. Alzheimers. Dis. 59, 893–901 (2017).

26. Bjørnarå, K. A., Dietrichs, E. & Toft, M. REM sleep behavior disorder in Parkinson’s disease--is there a gender difference? Parkinsonism Relat. Disord. 19, 120–122 (2013).

27. Bugalho, P., da Silva, J. A. & Neto, B. Clinical features associated with REM sleep behavior disorder symptoms in the early stages of Parkinson’s disease. J. Neurol. 258, 50–55 (2011).

